# The focus and timing of COVID-19 pandemic control measures under healthcare resource constraints

**DOI:** 10.1101/2020.04.16.20067611

**Authors:** Chen Wei, Zhengyang Wang, Zhichao Liang, Quanying Liu

## Abstract

Generalizing COVID-19 control strategies in one community to others is confounded by community’s unique demographic and socioeconomic attributes. Here we propose a tailored dynamic model accounting for community-specific transmission controls and medical resource availability. We trained the model using data from Wuhan and applied it to other countries. We show that isolating suspected cases is most effective in reducing transmission rate if the intervention starts early. Having more hospital beds provides leverage that diminishes with delayed intervention onset. The importance of transmission control in turn increases by 65% with a 7-day delay. Furthermore, prolonging outbreak duration by applying an intermediate, rather than strict, transmission control would not prevent hospital overload regardless of bed capacity, and would likely result in a high ratio (21% ∼ 84%) of the population being infected but not treated. The model could help different countries design control policies and gauge the severity of suppression failure.

## Introduction

Wuhan in China, being the first city to report cases of COVID-19, has seen 0 new cases for the first time on March 18, 2020 since the outbreak started in December 2019. Beyond implementing non-pharmaceutical interventions (NPI), including citywide traffic restrictions and social distancing to reduce contact rate^1^, the city of Wuhan further took proactive actions to isolate suspected cases in permanent and temporary medical facilities until laboratory tests to confirm such cases^1^. In contrast, some countries, such as US^2^ and Canada^3^, currently (March 2020) adopt alternative strategies preferring home-isolation for individuals showing symptoms, while only the severe cases are encouraged to be hospitalized, presumably as an attempt to prevent overloading the healthcare system.

Evaluating control policy efficacy requires an accurate and generalizable transmission model. Many early models regarding COIVD-19 transmission, however, were underinformed and missed certain critical assumptions. One of the earliest estimates of the number of the infected in Wuhan, China, based on the number of exported cases from Wuhan via international air travel, suggested a cumulative 4,000 symptomatic cases in Wuhan by January 18, 2020^4^. Adopting such methodology for early size estimation, Wu^5^ proposed a then influential susceptible–exposed–infected–recovered (SEIR) model by fixing the average period of infectivity at 2.4 days, which based on current evidence^6^, was too low and likely resulted in an over-estimation of the transmission rate. The study further assuming a flat 0%/50%/75% drop in transmissibility following Wuhan city lockdown. Transmission models with more sophisticated assumptions have since been proposed^7,8^, including ones with time-dependent control policies^9^ or non-Markovian transmission dynamics^10^. But these studies rarely made use of data beyond the confirmed number of cases such as the number of suspected cases. Another key factor that shall not be neglected in modelling is the real-time healthcare system capacity. While some studies have considered its impact^11^, few have incorporated such a factor dynamically in their transmission models.

The availability of a complete set of data and detailed records of adopted control policies from Wuhan enables us to construct and test an epidemic model that accounts for the factors deciding an outbreak profile, including the control policy’s evolution through time as well as the healthcare system capacity. We also dissected the effect of the proactive case isolation strategy adopted in Wuhan. The model validated with data from Wuhan is potentially applicable to other countries. Subsequently manipulating the parameters corresponding to control strategies opens a window to evaluate the effectiveness of each policy under certain healthcare system capacity constraints. We gauged the required NPI strength and timing for other 6 representative countries, namely US, Italy, France, Germany, Japan and Korea, with different hospital bed availability.

## Results

A modified SEIR model (Figure 1) with time-dependent transmission rate control, time-dependent case isolation and testing rate, non-Markovian patient discharge was proposed (see Methods for details). The model also imposes a hospital bed upper limit. Data from Wuhan between January 10, 2020 and March 11, 2020 was used to fit the model parameters.

**Figure 1.**
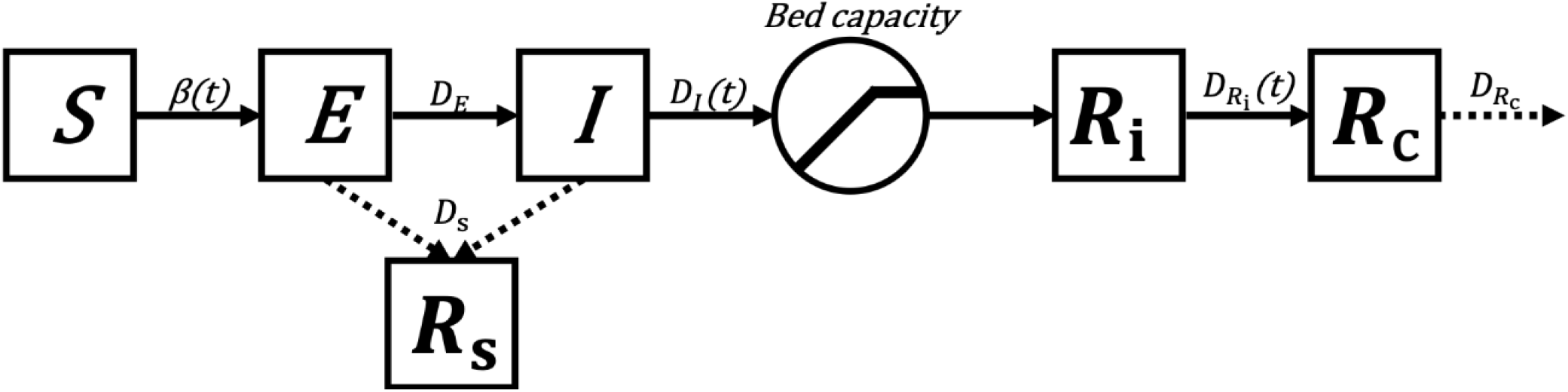
Modified SEIR model. Components include: *S* – susceptible cohort; *E* – asymptomatic cohort; *I* – symptomatic cohort; *R*_i_ – isolated cohort; *R*_s_ – the cohort losing infectivity without medical treatment; *R*_c_ – confirmed cohort. Bed capacity limited rate of isolation. Dashed arrows stand for cases leaving the dynamical system.

### Control Measures in Wuhan

As shown in Figure 2A, the combined effect of increases in NPI and case isolation explained the successful outbreak suppression in Wuhan. Specifically, the transmission rate in Wuhan was estimated to have been reduced to 31.0% of pre-lockdown level 5 days after January 23, 2020 and approaching an asymptote of 27.3%. Furthermore, the daily proportion of symptomatic cases being isolated in medical facilities was estimated to have increased from a starting estimate of 0.017 on January 10, 2020 to 0.081 on January 28, 2020. The daily proportion of isolated cases being confirmed was estimated to have started at 0.092 and increased to 0.22 by January 28, 2020. The basic reproduction number, corresponding to the initial stage of the outbreak, was estimated to be 3.48. Subsequently, the synergistic effect of increasing transmission control and isolation rate effectively reduced the daily reproduction number (see Suppression success in Methods) to be below 1 by Feb 28, 2020, subsequently resulting in a suppressed COVID-19 transmission (Figure 2B).

**Figure 2.**
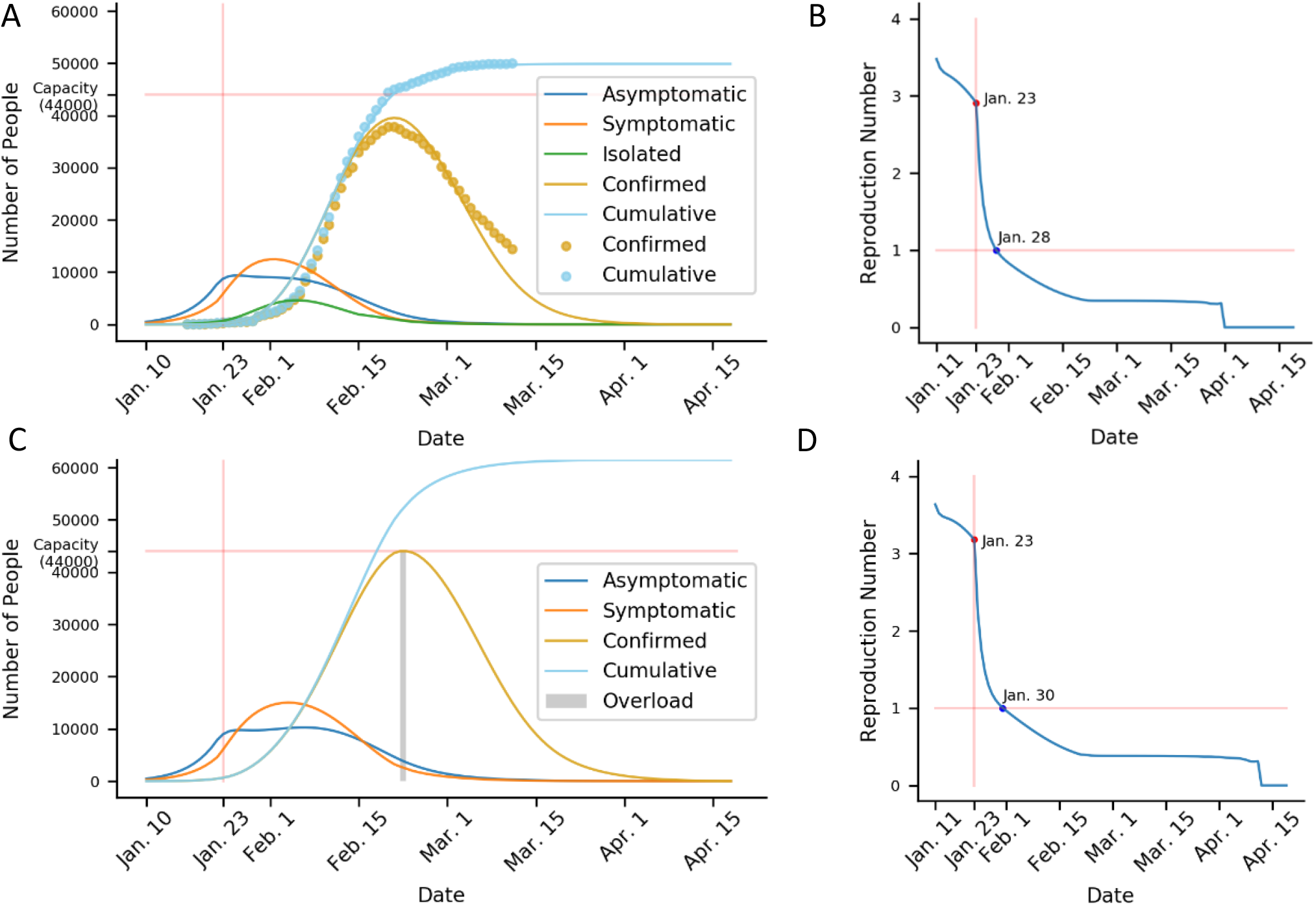
Modified transmission model fitting with Wuhan data. (A) Outputs of model fitted by the daily cumulative and current number of confirmed cases in Wuhan. Model outputs are plotted as solid lines and data as dots. The horizontal line stands for hospital bed capacity. (B) Evolution of the daily reproduction number with time in Wuhan. January 23 marks the onset of city lockdown. (C) Same as (A) but without proactive isolation. The shaded area indicates the duration of hospital overload. (D) Same as (B) but without proactive isolation.

The estimated number of early cases until January 18, 2020 combing both the symptomatic and asymptomatic was 5,101, which was in line with the number of 4,000 (95% CI [1,000, 9,700]) independently estimated from the number of COVID-19 cases exported from Wuhan internationally via air^4^. Throughout the outbreak, the highest bed occupancy attributed to COVID-19 (including both suspected and confirmed cases, see Methods) was estimated to be 40,710, which was below the reported maximum bed capacity of 44,000 in Wuhan.

Alternatively, we simulated that without proactive isolation, the daily reproduction number would have been reduced to 1 on January 30, 2020, 2 days later than reality (Figure 2D). In this case, Wuhan’s bed capacity would have been marginally saturated for 2 days between February 22,2020 and February 23, 2020 (Figure 2C). However, since the daily reproduction number would have been way below 1 at the time of hospital bed saturation, suppression would still have been achieved.

Through cross-validation (see Model evaluation in Methods), it was shown that the time-dependent model, which assumed exponentially decreasing transmission rate and isolation delay, had lower prediction error (mean ΔRMSE = -779.16, *p*(ΔRMSE < 0) = 0.758, Figure S2B), compared to the null model that assumed those factors to be constant. In addition, the time-dependent model would make increasingly better predictions than the null model for future time points with accumulating evidence (Figure S2A).

### Case isolation and policy timing

We further estimated the effects of onset timing of control policy on outbreak suppression. The utility of isolating suspected cases was evaluated based on its influence on the NPI strength threshold required for successful suppression. The criterion for a successful suppression was to reduce the daily reproduction number to 1 before all beds would have been occupied. As seen in Figure 3A, the threshold for successful suppression clearly defined the resulting number of total infections. The number of total infections exhibited a noticeably discontinuous jump around the suppression threshold. While successful suppressions would cause a small percentage (median = 0.46%) of the population to be infected, failed suppressions would lead to much more infections (median = 96.6%).

**Figure 3.**
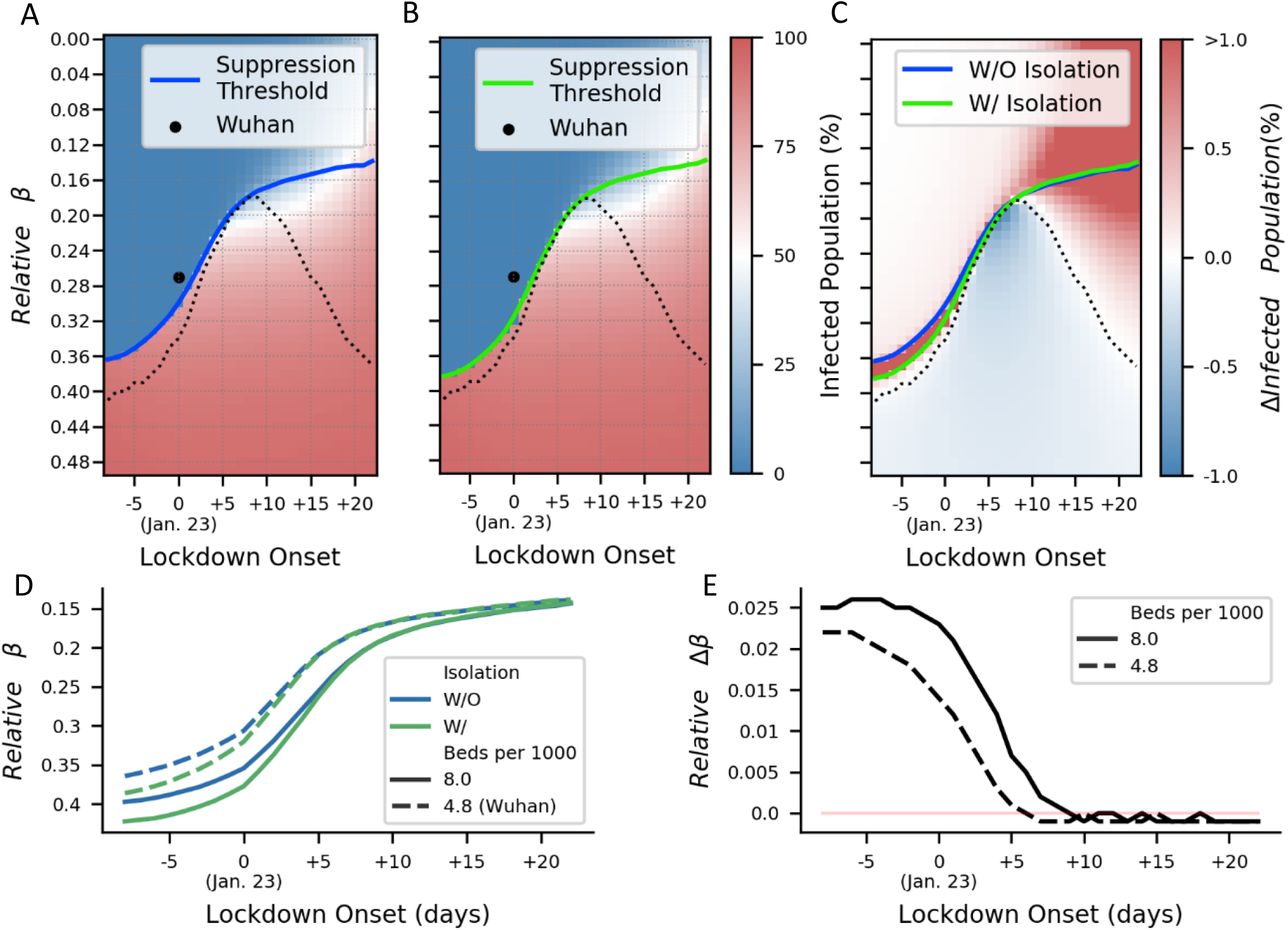
Case isolation effectiveness with different intervention timing. (A) Percentage of population that would have been infected in total with different relative transmission rate and lockdown onset date relative to January 23, 2020 without case isolation. The solid line indicates the suppression threshold according to the daily reproduction number. Onset date and fitted transmission rate was plotted for Wuhan. (B) Same as (A) but with proactive isolation. (C) Difference between (A) and (B). Not isolating suspected cases would have resulted in more infections under intervention designs represented by areas above the dashed line that also largely capture the scenarios of successful suppression. (D) Relative transmission rate thresholds assuming 4.8 and 8.0 beds per 1000 population at different intervention onsets. (E) Advantage of isolation in relative transmission rate threshold assuming 4.8 and 8.0 beds per 1000 population as a function of intervention onset time.

Naturally, the further delayed had the implementation of controls been, the harder it would have been to achieve suppression. Compared against the alternative strategy of only hospitalizing confirmed cases (Figure 3B), Wuhan’s strategy of hospitalizing both confirmed and suspected cases won a margin worth a 1.4% decrease in relative *β* requirement (Figure 3C&E). Nevertheless, fitting results showed that Wuhan’s NPI strength and timing would have been adequate to put the city well above the suppression threshold even without proactive isolation (Figure 3B). The benefit would have been the largest if actions had been taken as early as possible and would otherwise have been diminished by January 29, 2020, only 6 days later than the actual onset of intervention. Should suppression fail, however, the isolation strategy would have had a negative impact on the total number of infected cases. At most an extra 1.2% of the population would have been infected (onset = +6 days, relative *β* = 0.20) given proactive isolation (Figure 3C). Furthermore, increasing hospital bed limit from 4.8 beds (the real number in Wuhan) to 8.0 beds per 1,000 population would have provided a further cushion for transmission rate reduction, but such an advantage would also have been diminished if the intervention had been postponed for 9 days (Figure 3D&E).

### Leveraging hospital bed capacity in choosing NPI

Since most countries did not perform proactive isolations, the model without the isolation cohort was further examined. We manipulated hospital bed capacities estimated from their national averages, and searched for the suited transmission control measures for certain communities.

Both the duration of healthcare system overload and the final proportion of infected population were the measurements used to evaluate outbreak outcomes. The two measurements showed the same steep jump in value, indicating suppression failure (Figure 4A, B, D&E). This jump was well captured by the suppression threshold derived from the daily reproduction number (the blue curves in respective panels). Compared to the hospital capacity in US, the high hospital bed capacity in Germany would yield a higher threshold for suppression (Figure 4C, relative Δβ = 0.10). But such an advantage would be greatly reduced if interventions were delayed by a week (Figure 4F, relative Δβ = 0.065). By regressing the threshold-level relative *β* on the log-transformed number of beds per 1,000 population for a range of intervention onsets (Figure 5A), we found that hospital bed capacity would play the most important role with the intervention onset 2 days later than Wuhan. But having an advantage in hospital bed capacity would be less relevant if the onset was further delayed (Figure 5B). Specifically, the relative utility of transmission control in relation to bed capacity would be increased by 64.9% with a 7-day onset delay (slope shifted from 9.50 to 5.76).

**Figure 4.**
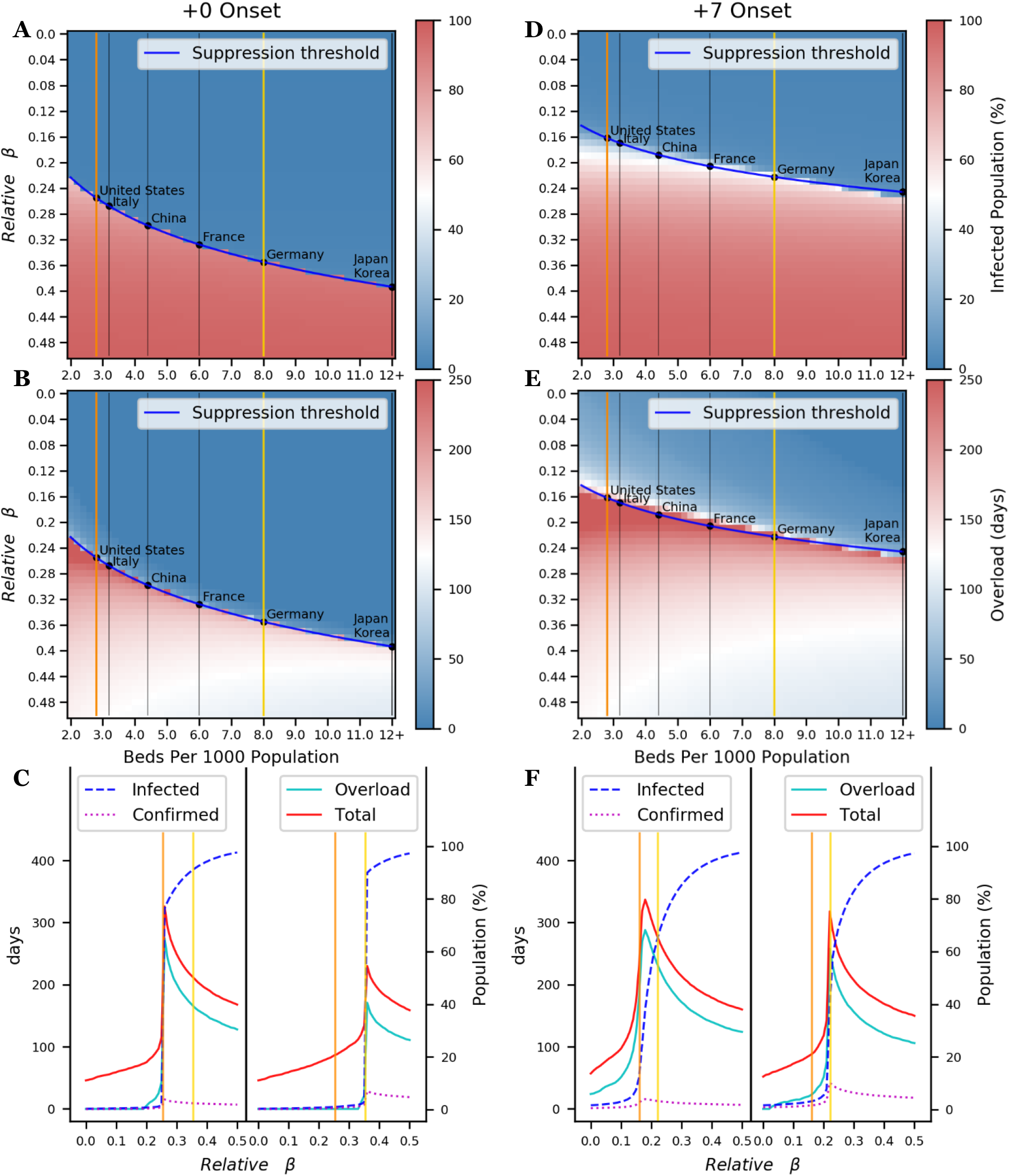
Suppression thresholds and failures. (A)Percentage of infected total population. The blue curve stands for the suppression thresholds. Its intersections with the vertical lines are the relative transmission rate thresholds for communities in exemplar countries with various bed capacities. The orange and yellow lines stand for the respective bed capacities of US and Germany. (B) Duration of hospital overload plotted in the same convention as (A). (C) Left: Percentage of total population infections, confirmed cases as a percentage of the total population, duration of outbreak and the duration of hospital overload for a community with bed capacity like that of US; Right: the same measures for a community with bed capacity like that of Germany. (D-F) The same as (A-C) except that the simulated intervention onsets were postponed by 7 days.

**Figure 5.**
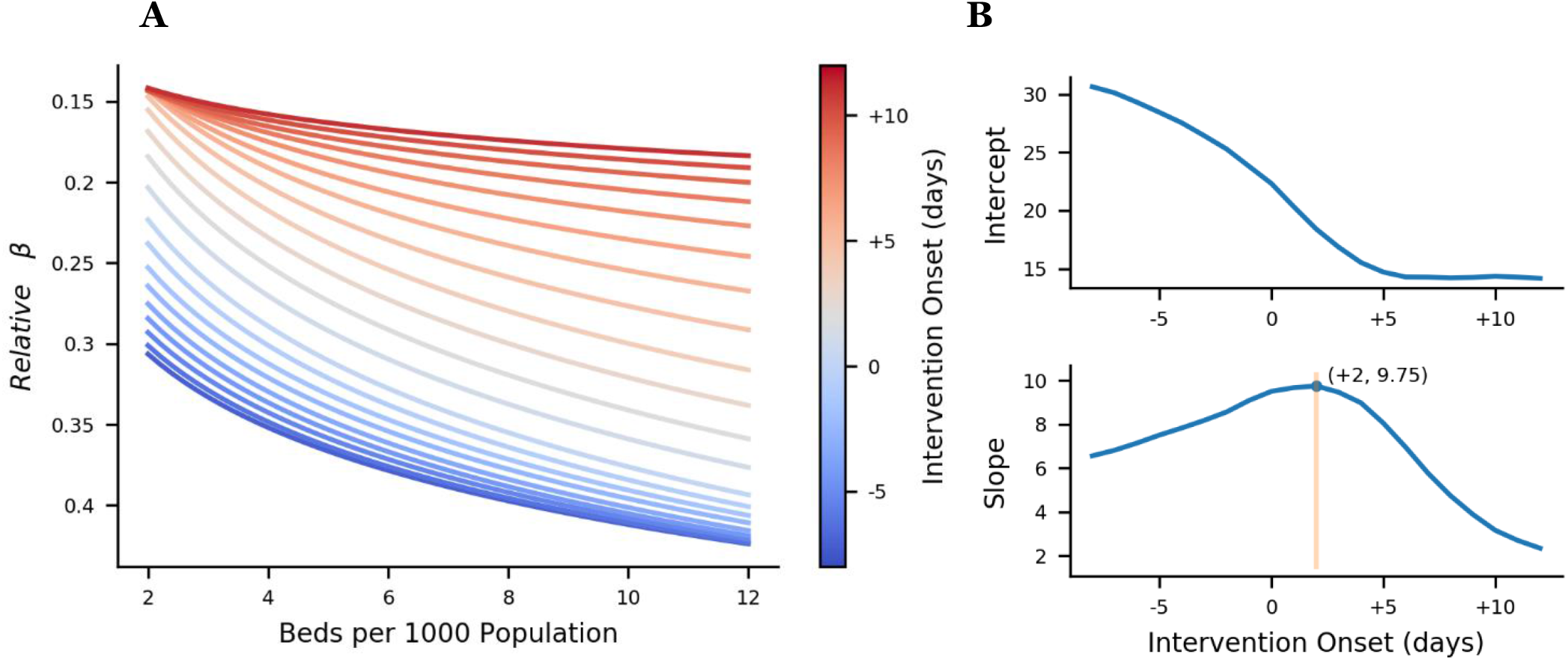
Suppression threshold as a function of relative transmission rate, beds per 1,000 population and intervention onset. (A) Logarithmic regression lines of beds per population on relative transmission rate. R^2^ range: [0.9147, 0.9961]. (B) Slopes and intercepts of the regressions in (A) for different intervention onsets.

The duration of hospital overload followed closely the total outbreak duration (+0 onset: correlation coefficient = 0.96; +7 onset: correlation coefficient = 0.99), with the maximum values of both observed near the suppression threshold (Figure 4C&F). Further relaxing transmission control (increasing relative *β*) from threshold value would cause the percentage of total infected population to steeply rise. The synergistic effect of extended hospital overload and infection number increase would lead to only a very small percentage of patients being treated.

In an attempt to evaluate the seriousness of suppression failure, we examined scenarios where relative *β* was at threshold level. A community with Germany level of healthcare capacity (8.0 beds per 1,000 population) and acting as promptly as Wuhan (0 days onset delay) would have a high relative *β* threshold. But if transmission control requirements were relaxed too much, it would still see 83.3% of the population infected but untreated (Figure 4C, right). To generalize, we simulated such scenarios with 0 to 7 days of onset delay and 2.8 to 8.0 beds per 1,000 population (see Figure S3). The results suggested that transmission control levels just short of suppression requirement, not only would yield the longest outbreak durations, but also would result in the size of untreated patients amounting to anywhere between 21.9% and 83.7% of total population.

## Discussion

Our model allowed retrospective analyses and counterfactual reasoning on Wuhan’s control policies. Wuhan’s strategy to proactively isolate all suspected cases, effectively removed patients from the infectious pool; otherwise waiting for confirmation would lead to heavier reliance on the laboratory testing capacity and thus leaving more infectious cases in the community. Serving as a buffer between the symptomatic cohort and the confirmed cohort, Wuhan’s strategy bought extra time for the culmination of medical resources in all departments, including extra-provincial medical staff dispatch, test kit production and new hospital construction. Our model showed that such a strategy in Wuhan was implemented soon enough (6 days until the advantage diminished) to provide a relieved NPI strength required for suppression. Moreover, proactive isolation in Wuhan was also beneficial in reducing peak hospital bed occupancy (from 44,000 to 40,710). As long as the transmission control had been strong enough, such positive outcomes would have been warranted regardless of the timing of NPI. However, if control measures had not been sufficiently strong to suppress the outbreak, isolating all suspected cases would have had a negative outcome due to imposing extra stress on the healthcare system.

More generally speaking, increasing NPI strength and hospital bed capacity both contribute to successfully suppressing the outbreak. However, the trade-off between investing in stronger NPI and larger healthcare system capacity depends on how promptly a community responds. As suggested in Figure 4, when the control measures are delayed and the early window is missed, not only does the overall difficulty to suppress become higher, advantages in healthcare system capacity also quickly diminish. Delaying intervention renders investing in NPI increasingly more efficient than boosting healthcare system capacity (Figure 5B).

Failing to suppress would cost countries even with the world’s more abundant hospital beds a steep increase in the number of untreated infections. Meanwhile, said communities would suffer an extended duration of healthcare system overflow. The “flattening the curve” notion was first illustrated by US Centers for Disease Control and Prevention (CDC)^12^ and later popularized on social media. It advocates for a level of transmission control that would be enough to lower the peak of patient number by prolonging the outbreak duration and thus preventing healthcare system overflow. However, our simulation proved this strategy to be flawed regarding COVID-19 (Figure 4). Successful suppressions are featured by greatly reduced outbreak size and duration. In contrast, prolonged outbreak durations indicate inadequate transmission control, in which case healthcare system overload is unavoidable (Figure 4C&F). Consequently, the number of untreated infections (and thus the death toll) would outgrow that of the treated by orders of magnitude (Figure 4F). Either consequence would be in violation of the goals behind the “flattening the curve” strategy. A similar conclusion has been drawn^11^ for the situations in UK and US to dissuade mere mitigation strategies. Nonetheless, if suppression is not achievable, the results still favor implementing as strict as possible transmission controls, since giving up transmission control in total would only further increase the number of untreated patients.

Instead of the explicit number of deaths from either untreated or treated patients, we paid attention to the number of infected patients that went untreated. In fact, this cohort could not be directly documented and thus are not directly reflected in the case numbers reported by countries. However, the significance of estimating those numbers became evident, as the simulation suggested that when suppression failed, the majority of the patients would be deprived of a chance at treatment (Figure 4 C&F). The exact mortality rate of COIVID-19 has been shown to be decided heavily by factors including patient age and pre-existing conditions^6,13–15^, and the contributions of these factors are still under much debate. Nonetheless, such measure helps demonstrate a crucial aspect of the pandemic’s impact on public health. It is mostly decided by NPI, regardless of the community-specific factors including demographic constituents and healthcare system preparedness^11,16^ (e.g. intensive care unit space and equipment availability) that would impact death toll as well as average patient prognosis.

The timing for lifting control policies and the potential for a rebound in transmission thereafter were not explicitly investigated in this study. Under a given situation of successful suppression where the majority of the population was not infected, herd immunity would have to be established by population-wide vaccination^11,17^. Until an effective vaccine is developed, a partial relaxation of transmission control policies while maintaining suppression would nonetheless be potentially achievable by ensuring a close monitoring of the daily reproduction number. The reason behind it was evident in Figure 3&4 where the daily reproduction number at the time of hospital overload proved to be a clear indicator of suppression success.

Giving up transmission control to achieve herd immunity might be adopted by some communities. Such a proposal could turn out even more ineffectual in the wake of the outbreak depending on SARS-CoV-2 immunity duration. Herd immunity achieves reproduction number reduction by removing individuals from the susceptible pool. Based on an estimated basic reproduction number of 3.48 by our model, 71% of the population would need to build and sustain immunity for SARS-CoV-2; otherwise, the reintroduction of the pathogen would cause the disease to circulate again. Another issue for herd immunity strategy is the duration of immunity. Although conclusions varied, evidence from SARS suggested a significant antibody reduction 2 years after infection^18–20^. In this case, longitudinal studies on this topic for COVID-19 would be crucial for designing vaccination strategies.

As much as it has been used to describe the intrinsic properties of COIVD-19, the basic reproduction number *R*_0_ is a function of the transmission rate *β*_0_ that depends that the frequency of human contact. An *R*_0_ estimated from Wuhan is thus most suited to be transferred to a community with a similar level of social interactions. This is especially true when trying to describe the early transmission dynamics when no controls are in place. Subsequent description of control policy strength using relative *β* introduces flexibility by accounting for all factors influencing transmission rate. Another caveat to the current study is that we neglected the inter-community traffic in the model, rendering the model most fitting to apply on a community on total lockdown. At the city level, discussions have started regarding locking down certain hot spots. At the country level, many have closed their borders to non-citizens. As the number of internationally imported cases grew, China announced that it would follow suit starting on March 28, 2020. Partial inter-city traffic reduction only has been estimated to be not effective in preventing inter-community spread^5,21^, thus the impact of travel on epidemic development through increasing both numbers of susceptible and infectious individuals would need to be carefully evaluated for future decision-making when reassessing lockdowns.

## Methods

### Data sourcing

Daily data from January 10, 2020 to March 11, 2020 including (a) the number of cumulative confirmed cases in Wuhan; (b) the number of currently hospitalized confirmed cases in Wuhan; (c) the number of new suspected cases in China; (d) the number of cumulative confirmed cases in China were obtained from the National Health Commission of the People’s Republic of China (http://en.nhc.gov.cn/). The number of daily new suspected cases in Wuhan was deduced from the number of daily new suspected cases in China based on the ratio of the cumulative number of cases reported in Wuhan to that reported in China.

Notably, the change in diagnostic criteria for COVID-19 on February 4, 2020, from solely relying on Polymerase chain reaction (PCR) to partially on clinical symptoms and chest CT scans, resulted in a single spike of 12,364 newly confirmed cases on February 12, 2020. By examining the evolution of the suspected case count on days preceding February 12, 2020, we concluded that the spike on February 12, 2020 was a delayed lump report of cases being cumulatively confirmed since the new criterion was implemented^22^. Thus, in order to better represent the historical trend of the testing capacity growth and maintain the cumulative count on February 12, 2020, the newly confirmed cases officially reported on February 12, 2020 were retrospectively assigned to 9 days between February 4, 2020 and February 12, 2020 by assuming a linear increase in daily confirmed cases through clinical diagnosis (Figure S1). Moreover, the number of available beds was estimated to be 44,000 according to the Health Commission of Wuhan website (http://wjw.wuhan.gov.cn/). The corresponding hospital bed capacities for other countries were based on the report^23^ by the Organisation for Economic Cooperation and Development (OECD).

### SEIR Model specification

A modified SEIR model was proposed and coded in Python. All citizens *N* are assumed to belong to the susceptible cohort *S*(*t*) at the beginning of the outbreak. Equation (1) asserts the susceptible to be infected by coming into contact with the asymptomatic cohort *E*(*t*) as well as the symptomatic cohort *I*(*t*) at a time-dependent rate of transmission *β*(*t*). The latent cohort *E* was set to be 50% as infectious, according to previous literature^24–29^. The asymptomatic cases are converted to the symptomatic, at an average rate of 1/*D*_*E*_ as seen in Equation (2) and Equation (3), where *D*_*E*_ is the incubation period and *D*_s_ is the period of infectivity. Furthermore, all population in the *E*(*t*) and *I*(*t*) cohort, without medical intervention, face a chance of spontaneously losing infectivity due to either recovery or COVID-19-related mortality, the average rate of which is define as 1/*D*_s_^6,30^. Removal from the *E*(*t*) population via such a route adequately accounts for the increasing evidence for asymptomatic yet infectious COVID-19 cases^24–29^.

Equation (4) and Equation (5) implement the process of isolation and confirmation. *R*_i_(*t*) represents the suspected and isolated case that were infected. Diagnosis precision p is computed as the ratio between the number of total suspected and confirmed cases and were used to adjust *R*_i_(*t*) to represent its actual contribution to bed occupancy. Cases in *R*_i_(*t*) were later confirmed through laboratory testing and became *R*_c_(*t*), the cohort of confirmed cases. *I*(*t*) flows into *R*_i_(*t*) at a rate of 1/*D*_*I*(*t*)_, while *R*_i_(*t*) flows into *R*_c_(*t*) at a rate of 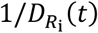. *D*_*I*(*t*)_ in Equation (7) specifies an exponential decrease in isolation delay starting on January 10, 2020 (*t*_0_), while the decrease in testing delay in Equation (8) follows the same exponential trend as that in isolation delay, reflecting a synchronization in resources utilized by either process (e.g. medical staff). The constraint of healthcare system capacity, an upper limit of *Bed*, limits the combine beds occupied by *R*_i_(*t*) and *R*_c_(*t*) in Equation (3) and Equation (4). The number of days a patient remains hospitalized until discharged are distributed uniformly amongst a 14-day time span centered at 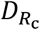, shown in Equation (5). The exponential decay of *β*(*t*) in Equation (6) describes the joint effect of the NPI measures from January 23, 2020 (*t*_1_) including traffic control, home-isolation, social distancing, disinfection and public use of personal protective equipment etc.

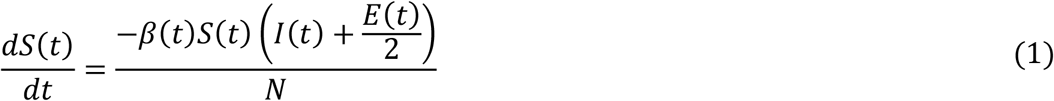

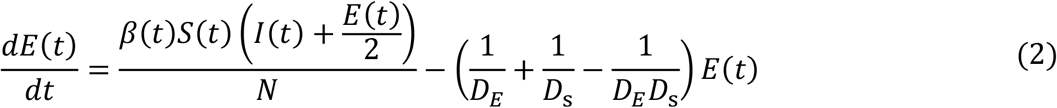

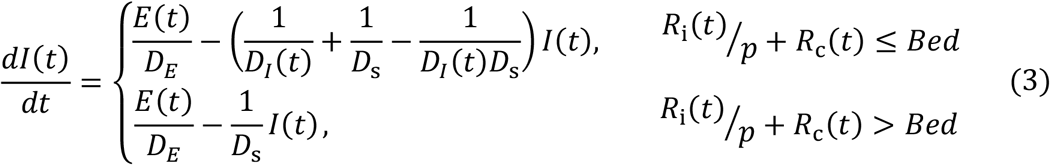

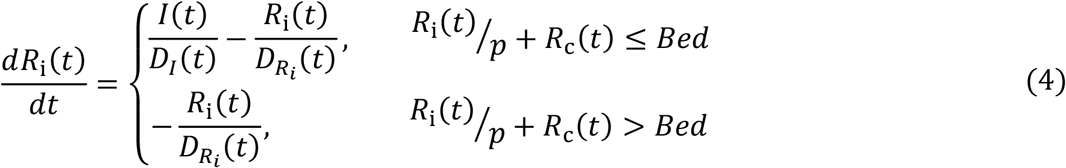

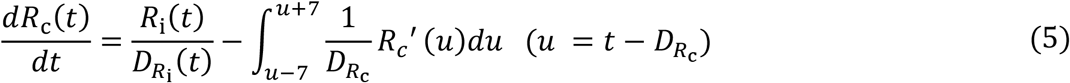

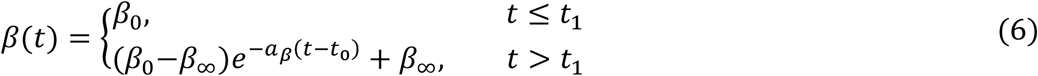

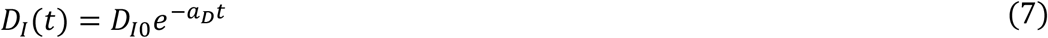

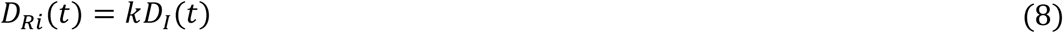

An alternative model assumes that the suspected patients are not isolated in medical facilities. Instead of flowing into *R*_i_(*t*), the suspected cases remain in *I*(*t*) until confirmed (Equation 9-10). The mean duration for a symptomatic case to be confirmed remains unchanged in Equation (11).

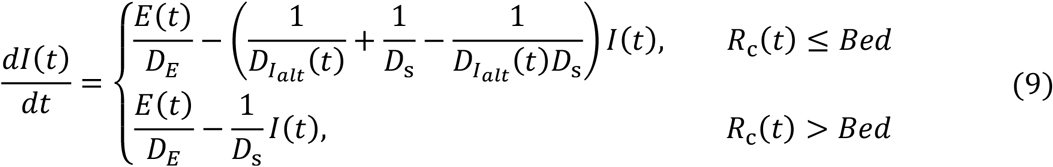

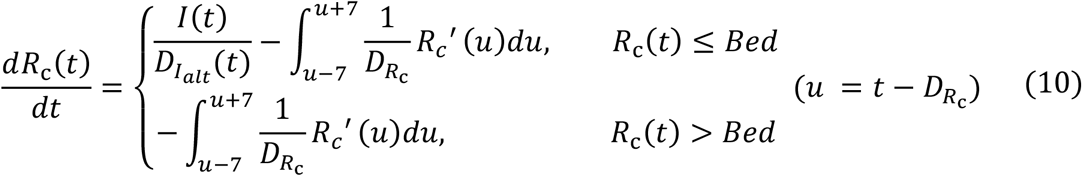

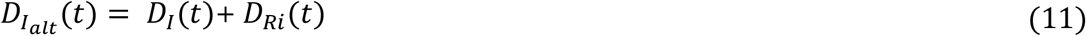

### Parameter estimation based on Wuhan data

The full profile of cumulative confirmed cases in Wuhan from January 10, 2020 to March 11, 2020 is used to estimate all free parameters except for 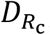, for these parameters only concern the inflow of infections while 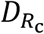 exclusively determines the outflow. A dual simulated annealing procedure is performed with 10,000 iterations using the *Scipy* package^31^ in Python. Subsequently, with the other parameters fixed, 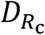 is estimated from the number of currently hospitalized confirmed cases in Wuhan using the same dual simulated annealing procedure. All parameters are summarized in Table 1.

**Table 1.** Parameters and their value in the model

| Parameter | Definition | Value | Source |
| --- | --- | --- | --- |
| $\beta_0$ | Initial transmission rate (proportion per day) | 0.692 | Dual simulated annealing |
| $a_\beta$ | Exponential decay constant of transmission rate | 0.5 | <sup>9</sup> |
| $\beta_\infty$ | Transmission rate asymptote (proportion per day) | 0.189 | Dual simulated annealing |
| $E(0)$ | Number of asymptomatic cases on January 10, 2020 | 318 | Dual simulated annealing |
| $I(0)$ | Number of symptomatic cases on January 10, 2020 | 227 | Dual simulated annealing |
| $1/D_{I0}$ | Initial isolation rate (proportion per day) | 0.0174 | Dual simulated annealing |
| $a_D$ | Exponential decay constant of isolation time constant | 0.0862 | Dual simulated annealing |
| $k$ | Ratio between isolation and confirmation rate | 0.391 | Dual simulated annealing |
| $D_{Rc}$ | Average delay of discharge or mortality (days) | 21.0 | Dual simulated annealing |
| $D_s$ | Period of infectivity (days) | 14.0 | <sup>6,30</sup> |
| $D_E$ | Incubation period (days) | 4.0 | <sup>13</sup> |
| $p$ | Diagnosis precision amongst suspected cases | 0.84 | Data (c) and (d) |

### Model evaluation

The time-dependent model is compared to a null model with a constant rate of transmission and a constant rate of isolation/confirmation. A cross validation method is applied to verify the improvement in prediction performance through a bootstrapping procedure described below^32,33^.

Data from January 15, 2020 to March 11, 2020, 56 days in total, are involved in the analysis. The first 28 days are regarded as the training set and the later as the test set. The model is trained by the data sampled from the training set, and then tested the data sampled from the test set. In this way, it can avoid the bias due to the testing data immediately follow the training data. To ensure sufficient data for training, the samples of training data vary from 14 to 28, whereas the samples of testing data span 1 to 28. Each model was optimized with the training set data. Root mean squared error (RMSE) in the predictions make by each model were then calculated on the testing set. In total, 120,000 sample pairs were drawn to construct distributions for prediction errors by the two models as well as that for the difference between the two errors.

### Suppression success

A successful suppression is judged based on the reproduction number. The reproduction number is defined as the average number of new infections each patient generates. When the reproduction number is less than 1, the number of new cases decreases. If the reproduction number is greater than 1, the number of new cases would increase until the susceptible pool runs out^34^. At the beginning of an outbreak when every individual is susceptible, the basic reproduction number *R*_0_ can by calculated from Equation (12), given that the asymptomatic cases are not infectious. With the current model specifying time-dependent parameters and 50% asymptomatic transmissibility, the average time of contact for the symptomatic and asymptomatic cohort is computed respectively in Equation (13) and Equation (14). A daily reproduction number is then computed as in Equation (17). Whether the average daily production numbers during healthcare system overload is below 1 is the criterion for successful suppression. The strength of NPI was quantified as the relative transmission rate *β* calculated from *β*_∞_/*β*_0_. The threshold level of relative *β* was linearly regressed on that of the log-transformed bed per 1,000 population to quantify the relative importance of the two.

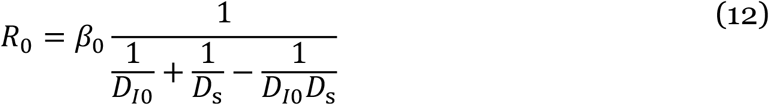

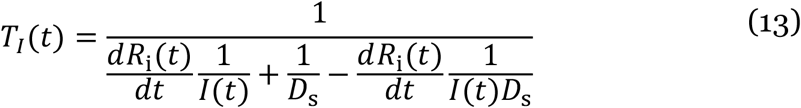

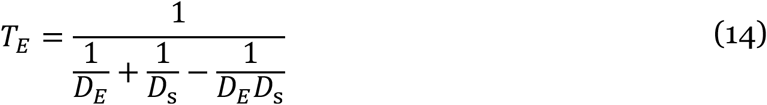

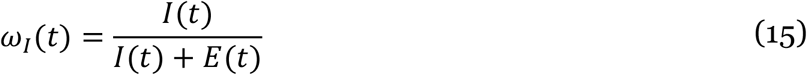

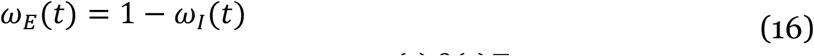

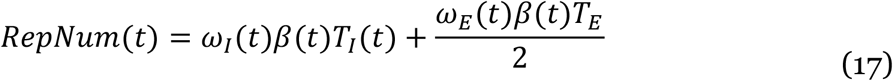

## Data Availability

The datasets generated during and/or analysed during the current study as well as the code used are available from the corresponding author on reasonable request.

## Acknowledgements

This work was supported by Guangdong Natural Science Foundation Joint Fund [No. 2019A1515111038] and High-level University Fund [No. G02386301, G02386401].

## Author contributions

C.W. performed analysis and wrote the paper. Z.W. performed analysis and wrote the paper. Z.L. performed analysis. Q.L. designed the research and wrote the paper.

## Supplementary information

## Simulation details

In Figure 3A&B&C, relative *β* increased from 0.0 to 0.5 (step=0.01); lockdown onset increased from -8 to 22 days (step = 1 day, Jan. 15-Feb. 15). The other parameters were kept fixed.

In Figure 4A&B&D&E, relative *β* increased from 0.0 to 0.5 (step=0.01) and number of beds per 1,000 population increased from 2.0 to 12.0 (step=0.2). Lockdown onsets were +0 and +7 respectively. In Figure 4C&F, the duration of outbreak was defined as the number of days from Jan 10 until the number of confirmed cases went to 0.

**Figure S1.**
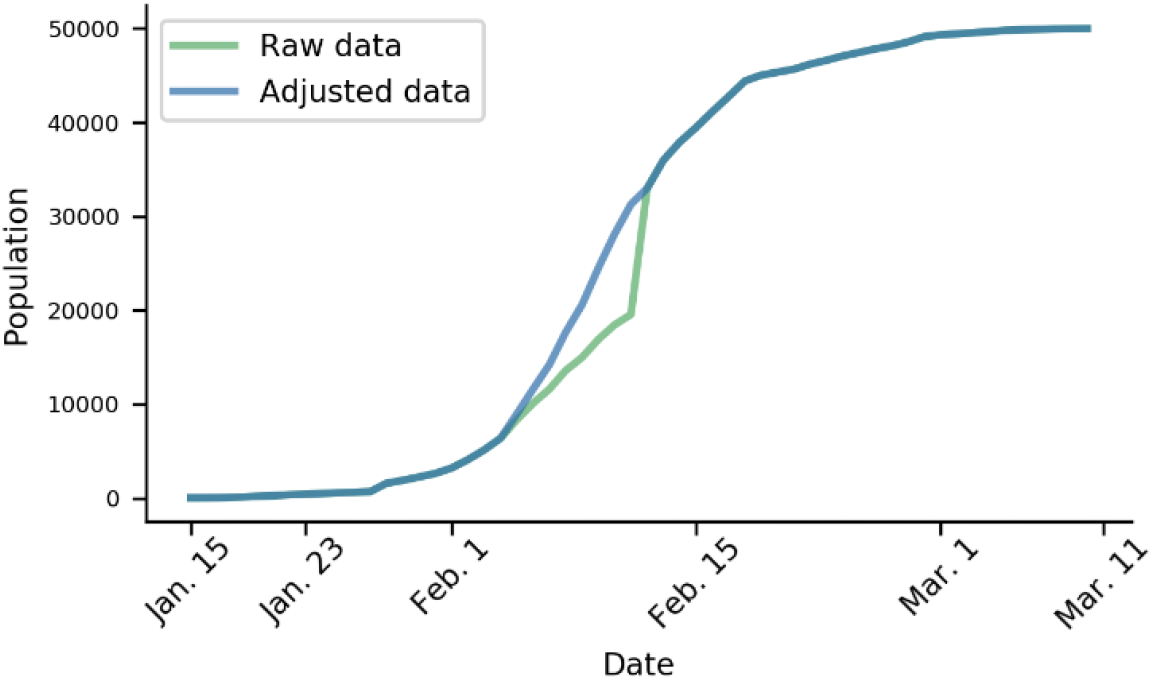
The cumulative confirmed data was adjusted to reflect updated diagnosis criteria.

**Figure S2.**
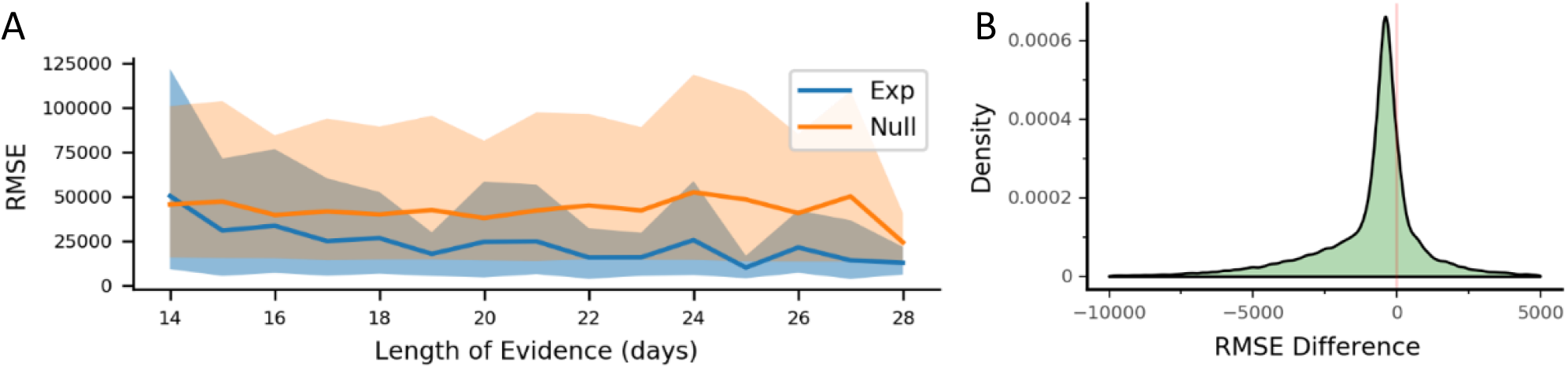
Model cross-validation. (A) Respective prediction errors as a function of evidence length by the model built with control-related parameters following exponential changes (Exp) and the null model with constant parameters (Null). Shades represent 95% quantile intervals. (B) Bootstrapped distribution of prediction error differences between the Exp and Null models using 1/4 to 1/2 of total data as evidence.

**Figure S3.**
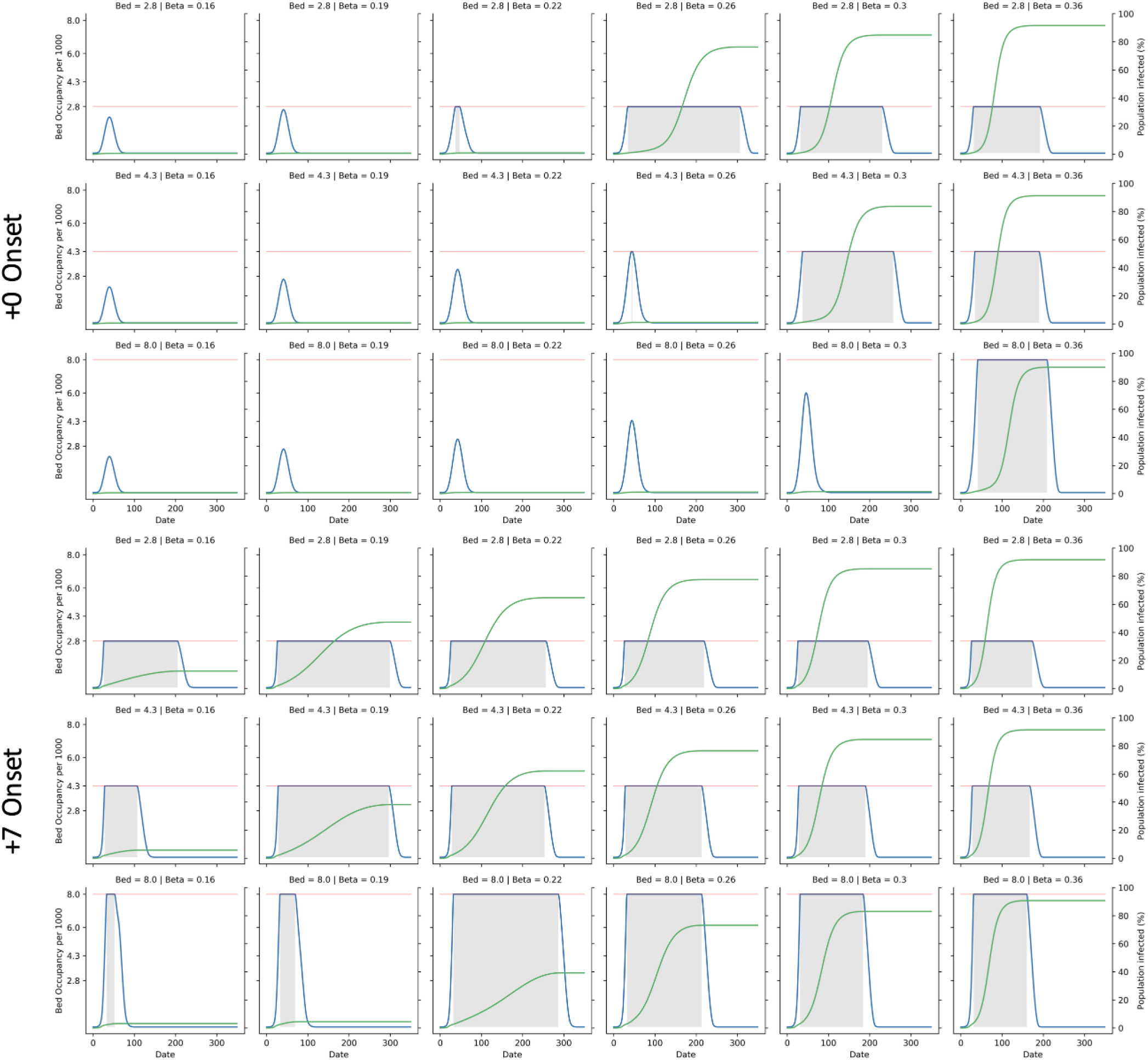
Evolution of hospital occupancy and infection count under different control policies. Each panel details one situation with the corresponding intervention timing, number of beds per 1,000 population and relative *β*. Blue lines indicate the number of occupied beds per 1,000 population and green lines indicate the percentage of infected population. Shaded regions are when beds are saturated.

## References

1. WHO. Report of the WHO-China Joint Mission on Coronavirus Disease 2019 (COVID-19). https://www.who.int/docs/default-source/coronaviruse/who-china-joint-mission-on-covid-19-final-report.pdf (2020).

2. CDC. Coronavirus Disease 2019 (COVID-19) – Prevention & Treatment. Centers for Disease Control and Prevention https://www.cdc.gov/coronavirus/2019-ncov/prepare/prevention.html (2020).

3. Canada, P. H. A. of. Coronavirus disease (COVID-19): Prevention and risks. aem https://www.canada.ca/en/public-health/services/diseases/2019-novel-coronavirus-infection/prevention-risks.html (2020).

4. Imai, N. et al. Estimating the potential total number of novel Coronavirus cases in Wuhan City, China. 6 (2020).

5. Wu, J. T., Leung, K. & Leung, G. M. Nowcasting and forecasting the potential domestic and international spread of the 2019-nCoV outbreak originating in Wuhan, China: a modelling study. The Lancet 395, 689–697 (2020).

6. Zhou, F. et al.. Clinical course and risk factors for mortality of adult inpatients with COVID-19 in Wuhan, China: a retrospective cohort study. The Lancet 395, 1054–1062 (2020).

7. Yang, Z. et al.. Modified SEIR and AI prediction of the epidemics trend of COVID-19 in China under public health interventions. J. Thorac. Dis. 12, (2020).

8. Lai, S. et al.. Effect of non-pharmaceutical interventions for containing the COVID-19 outbreak: an observational and modelling study. medRxiv 2020.03.03.20029843 (2020) doi:10.1101/2020.03.03.20029843.

9. Tang, B. et al.. An updated estimation of the risk of transmission of the novel coronavirus (2019-nCov). Infect. Dis. Model. 5, 248–255 (2020).

10. Chen, Y., Cheng, J., Jiang, Y. & Liu, K. A time delay dynamic system with external source for the local outbreak of 2019-nCoV. Appl. Anal. 0, 1–12 (2020).

11. Ferguson, N. et al.. Impact of non-pharmaceutical interventions (NPIs) to reduce COVID19 mortality and healthcare demand. http://spiral.imperial.ac.uk/handle/10044/1/77482 (2020) doi:https://doi.org/10.25561/77482.

12. Qualls, N. et al.. Community Mitigation Guidelines to Prevent Pandemic Influenza — United States, 2017. MMWR Recomm. Rep. 66, 1–34 (2017).

13. Guan, W. et al.. Clinical Characteristics of Coronavirus Disease 2019 in China. N. Engl. J. Med. (2020) doi:10.1056/NEJMoa2002032.

14. Ruan, Q., Yang, K., Wang, W., Jiang, L. & Song, J. Clinical predictors of mortality due to COVID-19 based on an analysis of data of 150 patients from Wuhan, China. Intensive Care Med. (2020) doi:10.1007/s00134-020-05991-x.

15. Wu, J. T. et al.. Estimating clinical severity of COVID-19 from the transmission dynamics in Wuhan, China. Nat. Med. 1–5 (2020) doi:10.1038/s41591-020-0822-7.

16. Ji, Y., Ma, Z., Peppelenbosch, M. P. & Pan, Q. Potential association between COVID-19 mortality and health-care resource availability. Lancet Glob. Health 8, e480 (2020).

17. Fine, P., Eames, K. & Heymann, D. L. “Herd Immunity”: A Rough Guide. Clin. Infect. Dis. 52, 911–916 (2011).

18. Wu, L.-P. et al.. Duration of Antibody Responses after Severe Acute Respiratory Syndrome. Emerg. Infect. Dis. 13, 1562–1564 (2007).

19. Liu, L. et al.. Longitudinal profiles of immunoglobulin G antibodies against severe acute respiratory syndrome coronavirus components and neutralizing activities in recovered patients. Scand. J. Infect. Dis. 43, 515–521 (2011).

20. Tang, F. et al.. Lack of Peripheral Memory B Cell Responses in Recovered Patients with Severe Acute Respiratory Syndrome: A Six-Year Follow-Up Study. J. Immunol. 186, 7264–7268 (2011).

21. Chinazzi, M. et al.. The effect of travel restrictions on the spread of the 2019 novel coronavirus (COVID-19) outbreak. Science (2020) doi:10.1126/science.aba9757.

22. 关于印发新型冠状病毒感染的肺炎诊疗方案 (试行第五版) 的通知. http://www.nhc.gov.cn/yzygj/s7653p/202002/3b09b894ac9b4204a79db5b8912d4440.shtml.

23. OCED. Hospital beds and discharge rates. in Health at a Glance 2019: OECD Indicators (OECD Publishing, 2019). doi:10.1787/0d67e02a-en.

24. Qiu, J. Covert coronavirus infections could be seeding new outbreaks. Nature (2020) doi:10.1038/d41586-020-00822-x.

25. Qian, G. et al.. A COVID-19 Transmission within a family cluster by presymptomatic infectors in China. Clin. Infect. Dis. Off. Publ. Infect. Dis. Soc. Am. (2020) doi:10.1093/cid/ciaa316.

26. Yu, P., Zhu, J., Zhang, Z., Han, Y. & Huang, L. A familial cluster of infection associated with the 2019 novel coronavirus indicating potential person-to-person transmission during the incubation period. J. Infect. Dis. (2020) doi:10.1093/infdis/jiaa077.

27. Rothe, C. et al.. Transmission of 2019-nCoV Infection from an Asymptomatic Contact in Germany. N. Engl. J. Med. 382, 970–971 (2020).

28. Bai, Y. et al.. Presumed Asymptomatic Carrier Transmission of COVID-19. JAMA (2020) doi:10.1001/jama.2020.2565.

29. Ganyani, T. et al.. Estimating the generation interval for COVID-19 based on symptom onset data. medRxiv 2020.03.05.20031815 (2020) doi:10.1101/2020.03.05.20031815.

30. Liu, Y. et al.. Viral dynamics in mild and severe cases of COVID-19. Lancet Infect. Dis. (2020) doi:10.1016/S1473-3099(20)30232-2.

31. Virtanen, P. et al.. SciPy 1.0: fundamental algorithms for scientific computing in Python. Nat. Methods 17, 261–272 (2020).

32. Racine, J. Consistent cross-validatory model-selection for dependent data: hv-block cross-validation. J. Econom. 99, 39–61 (2000).

33. Cerqueira, V., Torgo, L. & Mozetic, I. Evaluating time series forecasting models: An empirical study on performance estimation methods. ArXiv (2019).

34. Bailey, N. T. The mathematical theory of infectious diseases and its applications. (Charles Griffin & Company Ltd, 1975).

